# SARS-CoV-2 susceptibility and vulnerability of human blastocysts during *in vitro* preimplantation development

**DOI:** 10.64898/2026.11.08.26367551

**Authors:** Yana Kumpanenko, Maria Tryfonos, Wafaa Essahib, Neelke De Munck, Johan Van Weyenbergh, Yeranddy A. Alpizar, Kai Dallmeier, Hilde Van de Velde

## Abstract

Human preimplantation embryos express the SARS-CoV-2 entry factors ACE2, TMPRSS2 and BSG (CD147) on trophectoderm (TE) cells, suggesting susceptibility to infection. Likewise, SARS-CoV-2-pseudotyped reporter viruses induced cytopathic effects in blastocysts, while the zona pellucida (ZP) appeared to confer partial protection. The impact of authentic SARS-CoV-2 infection on embryonic lineage specification and developmental competence has not been established. Here, we exposed cryopreserved human embryos, donated under ethics approval and informed consent, to authentic SARS-CoV-2 (B.1.617.2, Delta variant). While embryos with an intact ZP remained uninfected, laser-assisted hatching rendered 19/21 (90.5%) of embryos susceptible to infection, as confirmed by detection of viral dsRNA. Moreover, SARS-CoV-2 infection had a pronounced effect on blastocyst development with a possibly fatal outcome. Compared to mock-treated embryos (*n* = 47), infected blastocysts (*n* = 48) showed a significantly reduced surface area (*p* = 0.0003), reduced total cell number (*p* = 0.0139), and reduced NANOG^+^ epiblast cell numbers (*p* = 0.0054) 24 hours post-exposure, while the number of GATA4^+^ primitive endoderm cells remained unaffected (*p* = 0.5255). Intriguingly, despite viral replication being largely confined to the trophectoderm (TE), this selective epiblast reduction suggests a disruption of TE-derived paracrine signaling required for epiblast specification. Finally, prolonged viral replication resulted in embryonic lysis and death. Digital transcriptomic profiling (mock, *n* = 29 embryos; SARS-CoV-2, *n* = 28 embryos; each in *n* = 5 minipools) revealed upregulation of innate antiviral and inflammatory genes, including *IFIH1*, *IRF7*, *NOD2*, and *TNFRSF1A*, as well as the recently identified SARS-CoV-2 entry factor *TMEM106B*, alongside altered developmental signatures concerning genome activation, pluripotency, and lineage-specific programs. These findings show that ZP disruption makes human blastocysts susceptible to productive SARS-CoV-2 infection, with adverse consequences for lineage allocation and direct implications for biosafety and virological counselling in medically assisted reproduction practice.

**Graphical Abstract:** 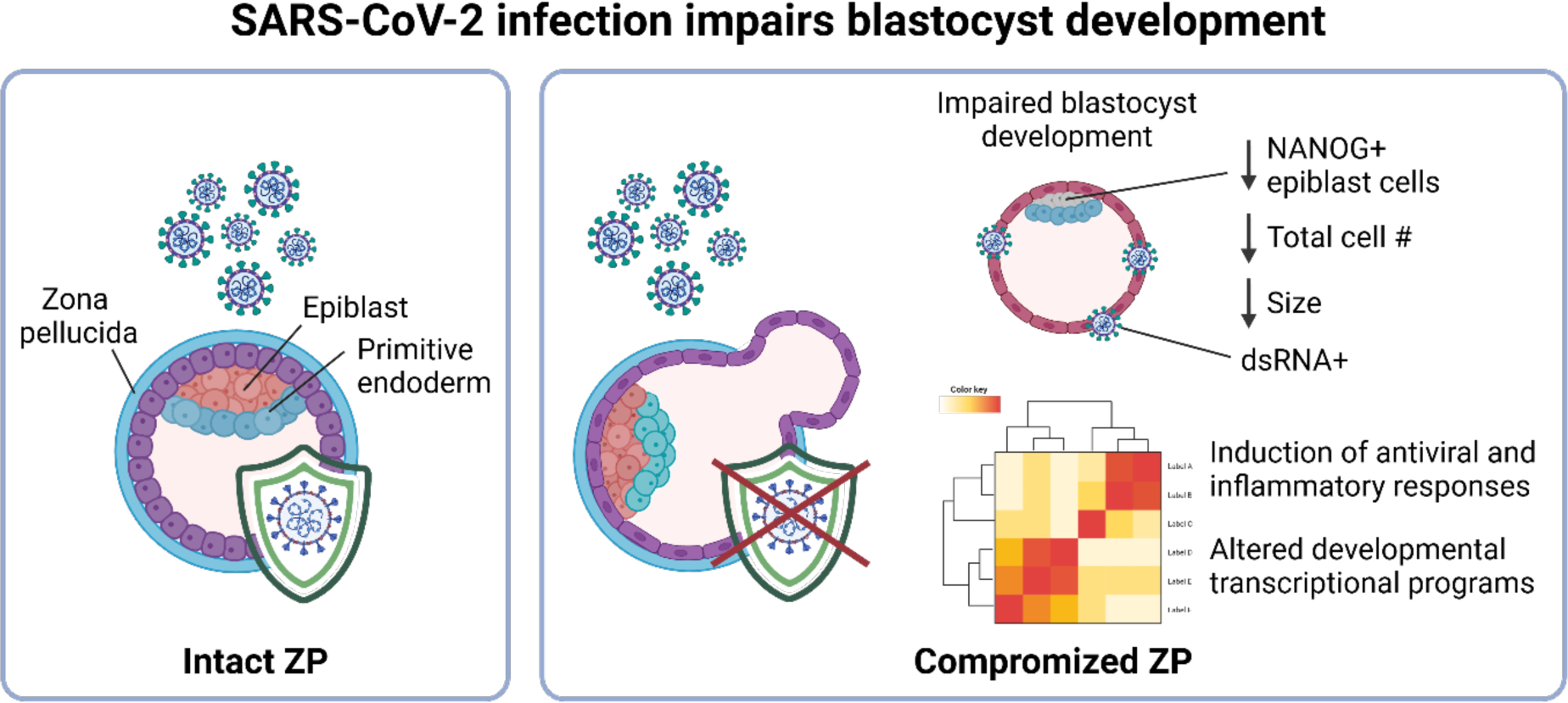

## INTRODUCTION

When SARS-CoV-2 appeared as a global health emergency in early 2020, its rapid spread posed an unprecedented challenge to healthcare systems worldwide, causing excess morbidity and mortality and disrupting medical care across virtually all specialties. Among the populations of particular concern were pregnant women and those undergoing medically assisted reproduction (MAR): clinicians and patients facing infertility treatment confronted an immediate and unanswered question: was it safe to continue^1^?

As clinical experience accumulated, evidence suggested that MAR technologies could generally be performed safely following recovery from SARS-CoV-2 infection, with clinical pregnancy, miscarriage, and live birth rates largely comparable to those of uninfected patients^2^. However, several studies reported reductions in embryo quality with molecular profiles of developmental delay, and an increase in miscarriage rate following recent infection, but the underlying mechanisms remain poorly understood^2–4^. Moreover, clinical outcome studies cannot determine whether these observations result from direct viral effects on the embryo or indirect maternal and endometrial factors, underscoring the need for experimental models that specifically address embryonic susceptibility to infection.

Beyond the preimplantation period, SARS-CoV-2 infection during pregnancy has been associated with an increased risk of adverse maternal and fetal outcomes, including preterm birth, fetal growth restriction, placental pathology, and stillbirth^5^. The severity of these complications differed between SARS-CoV-2 variants. Epidemiological studies consistently identified the Delta (B.1.617.2) variant of concern as being associated with the highest risk of severe maternal disease and adverse perinatal outcomes compared with earlier and subsequent variants^6,7^. Consequently, the Delta variant represents a clinically relevant model for investigating the effects of SARS-CoV-2 on early human development.

While these complications have been extensively investigated during established pregnancy, little is known about the effects of viral exposure before implantation, a question particularly relevant to MAR, where human gametes and embryos are manipulated outside the maternal environment and may be exposed to infectious agents during laboratory procedures. Although embryos are cultured under highly controlled conditions^8^, procedures such as oocyte retrieval, oocyte denudation (enzymatic softening and mechanical stripping of the cumulus and corona cells from an egg), embryo culture, assisted hatching, trophectoderm (TE) biopsy, and cryopreservation can transiently compromise their natural barriers.

Human embryonic development begins with fertilization, when a sperm cell fuses with an oocyte to form a totipotent zygote (day 1 post-fertilization, dpf)^9^. The embryo subsequently undergoes a series of cleavage divisions while enclosed within the zona pellucida (ZP), an extracellular glycoprotein matrix secreted during folliculogenesis^10^, reaching the compacted morula stage by approximately 4 dpf^11^. Around 5 dpf, cavitation gives rise to the blastocyst, composed of an outer TE and an inner cell mass (ICM)^12^. The ICM subsequently segregates into the NANOG-positive epiblast, which gives rise to the fetus, and the GATA4-positive primitive endoderm (hypoblast), which contributes to extraembryonic tissues^12,13^.

Throughout this period, the embryo remains enclosed within the ZP, which mediates species-specific sperm binding and prevents polyspermy, while protecting the conceptus from mechanical stress and enzymatic degradation during its transit through the fallopian tube^14,15^. Owing to its semi-permeable structure, the ZP also restricts the passage of large macromolecules and pathogens, including several viruses such as Adenovirus and SARS-CoV-2^16,17^. As the blastocyst expands, mechanical pressure combined with enzymatic degradation by TE-secreted and uterine proteases causes the ZP to thin and rupture, a process termed hatching, typically occurring at 5-6 dpf^18,19^. Hatching precedes implantation. It enables direct contact between the TE and the endometrial epithelium, thereby facilitating blastocyst apposition, adhesion, and subsequent invasion into the endometrium. However, hatching also marks the loss of the embryo’s protective shell, exposing the conceptus directly to the uterine environment for the first time, a vulnerability that becomes particularly relevant when considering exposure to infectious agents.

A molecular basis for embryo susceptibility was established when the entry receptors angiotensin-converting enzyme 2 (ACE2), Basigin (BSG/CD147), and transmembrane serine protease 2 (TMPRSS2), the principal host factors mediating SARS-CoV-2 cell entry, were detected on TE cells of human blastocysts, suggesting that preimplantation embryos possess the machinery required for viral infection^20,21^. However, viral RNA, measured by RT-PCR, has not been detected in follicular fluid, cumulus cells, or endometrial tissue of SARS-CoV-2-positive women^22^. Likewise, RT-PCR has detected viral RNA only very infrequently in the semen of infected males (pooled prevalence 1.76%)^23^. This suggests that the periconceptional environment may limit natural viral exposure at the gamete stage, although it does not exclude exposure, particularly in the laboratory during MAR.

A previous study showed that human blastocysts can be infected *in vitro* with SARS-CoV-2 spike-pseudotyped reporter virions; the resulting transduced embryos exhibited impaired developmental progression, and the study suggested that an intact ZP may protect against viral entry^17^. These findings provided the first evidence that preimplantation embryos might be susceptible to SARS-CoV-2 under experimental conditions.

While the above study modeled direct viral exposure *in vitro*, subsequent studies examined embryos derived from mothers with acute infection and found evidence of impaired development in more detail. Maternal infection prolonged the appearance of pronuclei, altered the expression of embryonic genome activation-related genes, and delayed the timing of the two-, three-, four-, and five-cell stages as well as blastulation. It was also associated with altered energy metabolism, enhanced glycolysis, and abnormal methylation/demethylation patterns in preimplantation embryos^4,24^.

However, these studies relied on modified viruses and largely qualitative assessments, and were limited to acute maternal infection, leaving several fundamental questions unresolved. It is still unclear whether authentic SARS-CoV-2 establishes productive infection and actively replicates within embryonic cells, whether infection differentially affects specific embryonic lineages, how profoundly it compromises blastocyst growth and cellular composition, and which molecular pathways are perturbed in response to infection. Addressing these questions is essential for understanding both the biological consequences of embryonic SARS-CoV-2 infection and the potential risks associated with viral exposure during the preimplantation period.

To address these questions, we investigated whether the SARS-CoV-2 Delta variant (B.1.617.2) can productively infect human preimplantation embryos and whether the integrity of the ZP determines susceptibility. We further examined the consequences of infection for blastocyst development, lineage specification, and transcriptional responses.

## MATERIALS AND METHODS

### Ethical statement

Human cryopreserved embryos were used following approval by the local ethical committee of UZ Leuven and UZ Brussel (BUN1432020000258) and the Belgian Federal Committee for research on human embryos (ADV_085bis_UZBrussel). Surplus cryopreserved embryos were donated to research, after informed written consent and 5 years of cryopreservation, by patients who had completed their fertility treatment at Brussels IVF (UZ Brussel) and no longer needed them.

### Culture of human embryos

Vitrified embryos were warmed and cultured at the research lab of Brussels IVF as previously described^25,26^. Briefly, 3 dpf (8-12 cell stage) embryos and 5 dpf (blastocyst stage) embryos, the latter derived from preimplantation genetic testing (PGT) cycles and therefore already possessing a ZP opened using LAH, were warmed using the Vit Kit-Thaw kit (90137-SO, Irvine Scientific) according to the manufacturer’s instructions. The 3 dpf embryos were cultured in equilibrated 25 μl droplets of blastocyst medium (83060010, ORIGIO Sequential Blast, with phenol red) overlaid with Ovoil (10174, Vitrolife) to prevent evaporation, at 37 °C, 5% O_2_, and 6% CO_2_ in a G185 flatbed incubator (K-Systems, CooperSurgical) for 24 h until 4 dpf before transport to the BSL-3 facility at the Rega Institute (KU Leuven). The 5 dpf embryos were allowed a 2-hour recovery period after warming before further transport. Only embryos that resumed development after cryopreservation were used in subsequent experiments; embryos showing signs of degeneration (including cytoplasmic vacuolization, cellular fragmentation, or cell lysis) in more than one cell were discarded. No clinical information on the donors, including maternal conditions or the indication for PGT, was available, and these factors could therefore not be accounted for in the analysis.

Following culture or recovery, embryos were transferred into microvials (5000-1020, Thermo Fisher Scientific) containing pre-equilibrated blastocyst medium and placed in a G95 Portable Incubator (K-Systems, CooperSurgical) at 37 °C for transport to the BSL-3 facility. There, embryos were immediately recovered from the microvials into fresh, pre-equilibrated 25 μl droplets under the same culture conditions. The 4 dpf embryos were cultured for a further 24 h until reaching the 5 dpf blastocyst stage before assignment to experimental groups; the 5 dpf embryos were randomly assigned to experimental groups directly following transport and re-equilibration. Embryos from the same experiment were warmed, cultured, and infected on the same day under identical conditions, so that each infected group had a matched mock group, and were subsequently allocated to downstream analysis by immunofluorescence or nCounter-based transcriptomic profiling; for the latter, embryos were transferred into 20 μl RLT lysis buffer (Qiagen) and snap-frozen.

### Opening/removal of zona pellucida

For the 3 dpf embryo groups, the ZP was opened by either laser-assisted hatching (LAH) or removed by chemical treatment with acid Tyrode’s solution (TS, pH 2.5 ± 0.3; T1788, Sigma-Aldrich).

LAH was performed as previously described^27^. Briefly, embryos were immobilized with the ZP positioned in the focal plane, and a series of consecutive laser pulses (80% intensity, millisecond-range pulse duration) was applied to a region distant from the blastomeres to create an opening of approximately 20 µm. Care was taken to avoid direct laser exposure of embryonic cells, after which the embryos were washed and returned to culture.

Chemical ZP removal was performed in 50 μl microdroplets of acid Tyrode’s solution or HEPES (4-(2-hydroxyethyl)-1-piperazineethanesulfonic acid; H6147, Sigma-Aldrich) medium supplemented with human serum albumin, under an Ivesta 3 Greenough stereomicroscope with integrated camera (Leica). Embryos were first rinsed in HEPES medium by gentle pipetting, then transferred to an acid droplet, where the outer zona layer was dissolved by gentle pipetting under continuous observation. Once the outer layer was nearly fully dissolved, pipetting was stopped immediately to avoid damaging the embryo. Embryos were then washed three times in HEPES medium and transferred to a new culture dish.

### SARS-CoV-2 viral infection

All work involving live virus was conducted in the high-containment BSL-3 facilities of the KU Leuven Rega Institute under license SBB219.2024/1599, in accordance with institutional guidelines.

SARS-CoV-2 Delta strain (B.1.617.2; EPI_ISL_2425097) virus stock was propagated in Vero E6 cells, and the median tissue culture infectious dose (TCID_50_) was determined by end-point titration using A549-Dual hACE2-TMPRSS2 cells (Invivogen, Cat. No. a549d-cov2r), as previously described^28^.

For the infection assay, blastocyst-stage embryos were placed in equilibrated 25 μl droplets of blastocyst medium mixed either with Dulbecco’s Modified Eagle Medium (DMEM, 10564011, Gibco) supplemented with fetal bovine serum (2%, A5256701, Gibco) as a control, or with virus at concentrations of 3×10^3^, 3×10^4^, and 3×10^5^ TCID_50_/ml. Embryos were cultured at 37 °C, 5% O_2_, and 6% CO_2_ for 24 h, until 6 dpf.

### Immunofluorescence

Embryos were fixed in 4% paraformaldehyde (047392.9L, Thermo Scientific Chemicals) in 1X PBS (10010023, Gibco) for 10 min at room temperature (RT) to cross-link proteins and preserve cellular integrity. After fixation, embryos were washed in 2% BSA (A2153, Sigma-Aldrich) in 1X PBS for 5 min, then permeabilized in 0.1% Triton™ X-100 (×100, Sigma-Aldrich) in 1X PBS for 20 min at RT with gentle side-to-side agitation on a reciprocal shaker at 100 rpm. To reduce non-specific antibody binding, embryos were transferred to a blocking buffer of 10% FBS and 2% BSA in 1X PBS and incubated for 1 h at RT. Embryos were then incubated overnight at 4 °C in primary antibody (**Table 1**) diluted in 10% FBS-2% BSA in 1X PBS. Following overnight incubation, embryos were washed in 2% BSA-1X PBS (3 × 10 min) and incubated for 1 h in secondary antibody (**Table 1**) diluted in 10% FBS-2% BSA in 1X PBS. From this step onward, embryos were kept in the dark at RT with agitation. For nuclear staining, embryos were further incubated for 20 min in Hoechst 33342 (H3570, Invitrogen™) diluted in 10% FBS-2% BSA in 1X PBS. Finally, embryos were washed in 2% BSA in 1X PBS (3 × 5 min) and transferred to 10 μl droplets of 2% BSA in 1X PBS on an Ibidi μ-Slide 8-well (80826, Ibidi) for imaging.

**Table 1.** Primary and secondary antibodies used in immunolabeling.

| Primary antibody | Host / Isotype | Dilution | Catalog # |
|---|---|---|---|
| Anti-NANOG | Mouse / IgG1 monoclonal NNG-811 | 1:50 | Abcam, ab21624 |
| Anti-GATA4 | Rat / IgG2a, kappa Monoclonal eBioEvan | 1:500 | Invitrogen, 4998082 |
| Anti-ACE2 | Rabbit / Polyclonal IgG | 1:1000 | Abcam, ab15348 |
| Anti-dsRNA (J2) | Mouse / IgG2a kappa monoclonal | 1:250 | SCICONS, 10010200 |
| **Secondary antibody** | **Host** | **Dilution** | **Catalog #** |
| anti-Mouse IgG2a Alexa Fluor™ 647 | Goat | 1:200 | Invitrogen, A21241 |
| anti-Rabbit IgG (H+L) Alexa Fluor™ 647 | Donkey | 1:200 | Invitrogen, A31573 |
| anti-Mouse IgG (H+L) Alexa Fluor™ 594 | Donkey | 1:200 | Invitrogen, A21203 |
| anti-Rat IgG (H+L) Alexa Fluor™ 488 | Donkey | 1:200 | Invitrogen, A-21208 |

Embryos were imaged using a confocal microscope (LSM800 equipped with Airyscan, ZEISS) at 40X magnification. Z-stack scans were acquired for whole-embryo visualization and 3D reconstruction. Images were analyzed in FIJI^29^. Each channel was manually adjusted for brightness and contrast, with the same settings applied across all images of a given stain within each experiment. The number of cells belonging to each embryonic lineage (NANOG for epiblast, GATA4 for primitive endoderm) was assessed manually; a nucleus was considered positive if its signal was visibly above background.

### Embryo size measurement

Embryo size was estimated from images taken at [0, 18, and 24 hpi] (Leica S9i brightfield images). In Fiji, the vertical and horizontal diameters of each embryo were measured, and the surface (S) was estimated as S = vertical diameter x horizontal diameter [mm^2^].

### Digital transcriptomics (nCounter)

Digital transcriptomics (nCounter platform, Nanostring Ltd.) was performed as previously described for other viral pathologies and their vaccine outcomes^30–32^. nCounter counts transcripts directly by hybridization to target-specific probes, without reverse transcription or amplification, which makes it suitable for very low RNA inputs. In brief, embryo lysates were hybridized to unique capture/reporter pairs (50 bp each) targeting 770 transcripts (730 immune transcripts and 40 internal controls present in the “Human Myeloid Innate Immunity” Panel V2, as well as virological transcripts (SARS-CoV-2 transcripts and ACE2/TMPRSS2 receptors), 6 positive and 8 negative control probes (all from NanoString). Results were sequentially corrected for background (negative control probes), technical variation (positive control probes), and RNA content (housekeeping genes).

A single blastocyst does not contain enough RNA to be reliably detected on this platform, so 5-6 embryos were pooled. Pool sizes were kept as equal as possible between mock and infected groups. Phenotypic readouts were assessed per embryo, whereas transcriptomic data represent pool-level measurements.

Counts were corrected sequentially for background (negative control probes), technical variation (positive control probes), and RNA content (housekeeping genes). nCounter is a targeted method: the panel focuses on innate immunity and was not designed for embryonic development, so genes relevant to early development may be missing, and novel transcripts cannot be identified. The transcriptomic findings are therefore considered exploratory.

### nCounter gene-expression and module analysis

Each nCounter sample corresponded to a pool of five to six embryos from a single group and experiment (*see Digital transcriptomics*). Housekeeping gene-normalized counts were log2-transformed after addition of a pseudocount of 1, and genes with zero variance across samples were excluded from statistical analyses. Differential expression between SARS-CoV-2-treated and matched mock-treated pools from the same experiment was analyzed using limma, with experiment and treatment included in the linear model to account for the paired design. Empirical Bayes moderation was applied using robust variance estimation and mean-variance trend modelling. The SARS-CoV-2-versus-mock contrast was used to calculate log2 fold changes, moderated *t*-statistics, and nominal *p* values, and *p* values were adjusted for multiple testing using the Benjamini-Hochberg method. Differentially expressed genes were defined by an absolute log2 fold change of at least 1 (a two-fold change in expression) and a false discovery rate (FDR) < 0.05.

Principal component analysis was performed on log2-transformed expression values of all non-constant genes, without variance scaling. For visualization, expression values were standardized separately for each gene across samples and displayed as gene-wise z-scores.

Predefined transcriptional modules were obtained from a previously published study^12^. For each module, only genes that were both included in the published module definition and represented in the nCounter panel were retained for analysis (**Tables 2 and 3**). Module scores were calculated as the mean z-score of all non-constant genes assigned to that module, and modules with fewer than two informative genes were excluded. Module scores were compared between matched SARS-CoV-2- and mock-treated pools using two-sided paired *t* tests, followed by Benjamini-Hochberg correction across modules. All analyses were performed in R using a custom script.

**Table 2.** Gene composition of transcriptional modules used for analysis. Genes were selected from the module definitions described in^12^ and restricted to those represented in the nCounter panel.

| **Module** | **Genes listed in<sup>12</sup>** | **Genes matched in nCounter panel** |
|---|---|---|
| **ZSCAN4** | 1480 | 55 |
| **DUXA** | 567 | 10 |
| **POU5F1B** | 149 | 8 |
| **DNMT3L** | 261 | 8 |
| **IFI16** | 144 | 16 |
| **GATA4** | 18 | 4 |
| **GATA2** | 335 | 21 |
| **NR2F2** | 536 | 41 |

**Table 3.**
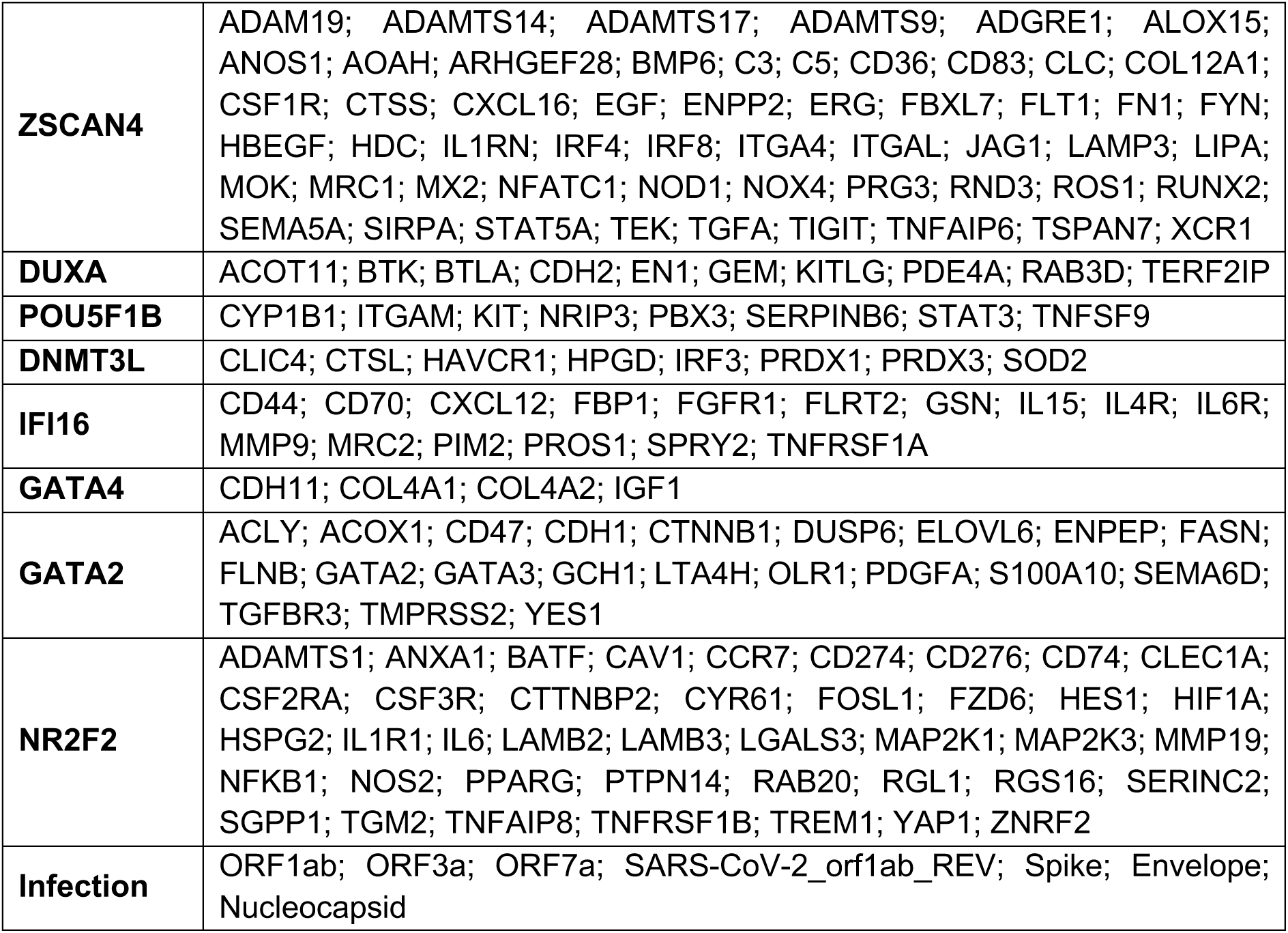
List of genes per module.

### Statistics

Statistical analyses and data visualization were performed using GraphPad Prism version 10 (GraphPad Software, San Diego, CA, USA). Differences between groups were assessed using Fisher’s exact test, multiple unpaired *t* tests, Mann-Whitney *U* tests, or ratio-paired *t* tests, as appropriate. All tests were two-sided, and *p* < 0.05 was considered statistically significant. Sample sizes (*n*), exact *p* values, and details of the statistical tests used are provided in the respective figure legends. For nCounter data, the unit of analysis was the embryo pool, and each SARS-CoV-2 pool was paired with a mock pool from the same experiment; for imaging data (surface area, cell counts, dsRNA staining), the unit of analysis was the individual embryo.

## RESULTS

### Laser-assisted hatching establishes a robust model for SARS-CoV-2 infection of human blastocysts

To establish a reproducible model for SARS-CoV-2 infection of human preimplantation embryos, we first compared two approaches to compromise the ZP: laser-assisted hatching (LAH) to create an opening in the ZP and treatment with Tyrode’s acid solution (TS) to dissolve the ZP (**Figure 1A**). Since it was previously shown that intact ZP might restrict efficient viral entry, ZP breaching is a prerequisite for establishing a reliable infection model^17^.

**Figure 1.**
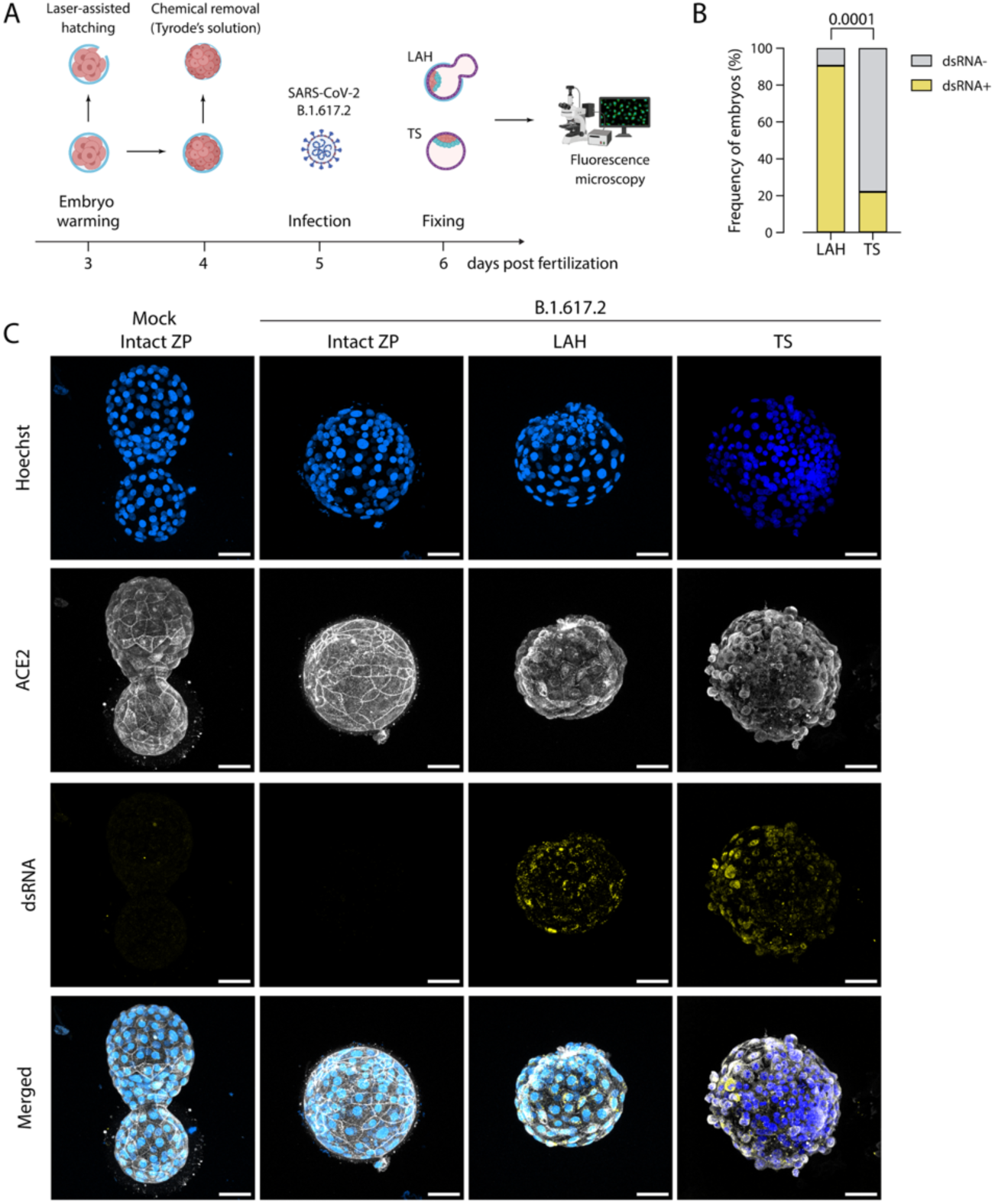
Laser-assisted hatching establishes a robust model for SARS-CoV-2 infection of human preimplantation embryos. **A** Experimental workflow. Vitrified human embryos (3 dpf) were warmed and cultured. Zona pellucida (ZP) compromise was achieved either by laser-assisted hatching (LAH) on 3 dpf or treatment with Tyrode’s acid solution (TS) on 4 dpf. A subset of embryos was maintained with an intact ZP. Embryos were cultured to 5 dpf and exposed to SARS-CoV-2 (B.1.617.2, Delta variant) or mock medium before fixation at 24 hpi. **B** Percentage of embryos positive for double-stranded RNA (dsRNA) following infection. LAH group, *n* = 21; TS group, *n* = 27. Fisher’s exact test, *p* < 0.0001. **C** Representative immunofluorescence images of mock, ZP-intact, LAH, and TS-treated embryos following SARS-CoV-2 exposure. Viral replication was detected by the presence of dsRNA (yellow). Nuclei were stained with Hoechst (blue). Expression of ACE2 receptor (white) is shown for reference. Scale bars, 50 μm.

Donated 3 dpf embryos were warmed at IVF Brussel and assigned to one of three groups. In the LAH group, partial ZP ablation was performed on the day of warming after embryo recovery. In the untreated group, the ZP was left intact throughout to assess susceptibility to infection without ZP manipulation. In the TS group, the ZP likewise remained intact at this stage. All three groups were cultured to 4 dpf and transported to the BSL-3 facility, where TS-designated embryos underwent gentle chemical ZP removal following a 2-hour recovery period. Culture then continued to 5 dpf, when embryos were randomly assigned to SARS-CoV-2 or mock infection. Untreated, ZP-intact embryos were challenged with 3×10⁵ TCID_50_/ml of the SARS-CoV-2 (Delta) variant, whereas LAH- and TS-treated embryos were infected with 3×10⁴ TCID_50_/ml. All embryos were fixed 24 hours post-infection (hpi) for immunofluorescence analysis.

LAH consistently generated embryos susceptible to productive infection. Double-stranded RNA (dsRNA), detected by J2 immunostaining as a marker of active viral replication^33^, was present in 19 of 21 LAH-treated embryos (90.5%), compared with only 6 of 27 TS-treated embryos (22.2%; Fisher’s exact test, *p* < 0.0001) at the viral dose used for quantification (3×10^3^ TCID_50_/ml) (**Figure 1B**).

In contrast, embryos with a completely intact ZP remained dsRNA-negative despite exposure to a ten-fold higher viral inoculum (3×10^5^ TCID_50_/ml), demonstrating that an intact ZP efficiently prevents SARS-CoV-2 infection. Consistent with this, partially hatched embryos showed dsRNA signal restricted to the exposed TE cells with no signal detected in the region still covered by ZP, indicating that the ZP acts as a local physical barrier to infection rather than conferring protection at the whole-embryo level (**Supplementary Figure 1A**).

Interestingly, one embryo with an apparently intact ZP displayed a dsRNA-positive signal, localized to a small, discrete spot rather than distributed across the embryo, with signal confined to the cytoplasm of approximately two TE cells (**Supplementary Figure 1B**). As this embryo originated from intracytoplasmic sperm injection (ICSI), the infection may have occurred through the microscopic opening created during sperm injection.

TS-treated embryos also exhibited reduced tolerance to viral exposure. At 3×10^4^ TCID_50_/ml, the dose used for LAH embryos, 9 of 13 TS-treated embryos degenerated following infection (**Supplementary Figure 1C**). To preserve embryo viability, the viral inoculum for the TS group was therefore reduced to 3×10^3^ TCID_50_/ml. The 22.2% infection rate and all subsequent analyses of TS-treated embryos are based on this lower viral dose.

Representative immunofluorescence images illustrate the marked difference in infection efficiency between intact and removed ZP (**Figure 1C**). dsRNA signal was distributed throughout the TE in the LAH and TS groups, with localized patches of stronger signal. Together, these findings demonstrate that efficient disruption of the ZP is essential for productive SARS-CoV-2 infection of human blastocysts. Compared with TS treatment, LAH provided substantially higher infection efficiency while maintaining embryo viability, establishing it as the preferred method for generating a robust and reproducible embryo infection model.

### SARS-CoV-2 infection reduces blastocyst growth, selectively affecting the epiblast lineage

Following establishment of the infection model, we investigated the effect of SARS-CoV-2 infection on blastocyst development using donated 5 dpf blastocysts that had previously undergone TE biopsy for preimplantation genetic testing (PGT) (**Figure 2A**). As these embryos already had a laser-created opening in the ZP, no additional manipulation was required before infection. After warming, embryos were transported to the BSL-3 facility, allowed to recover for 2 h, and either mock-infected or infected with the SARS-CoV-2 B.1.617.2 (Delta) variant (3×10^4^ TCID_50_/ml). At 24 hpi, embryos were fixed for immunofluorescence analysis or processed for digital transcriptomic profiling using the NanoString nCounter platform.

**Figure 2.**
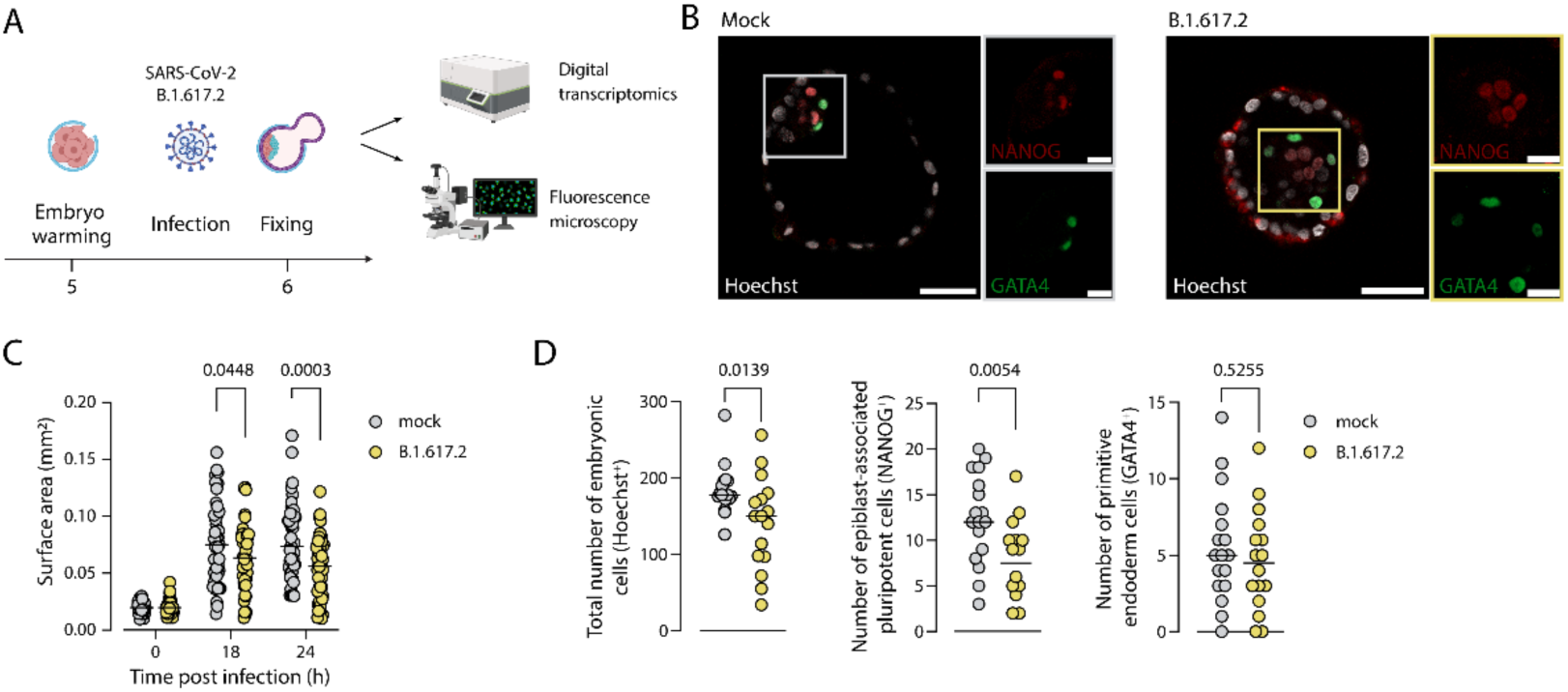
SARS-CoV-2 infection impairs blastocyst expansion and reduces epiblast cell numbers. **A** Experimental design. Blastocysts (5 dpf) were warmed, transported to the BSL-3 facility, allowed to recover for 2 hours, and mock-infected or infected with SARS-CoV-2 (B.1.617.2, Delta variant). At 24 hpi, embryos were processed either for immunofluorescence analysis or NanoString nCounter digital transcriptomic profiling. **B** Representative immunofluorescence images of mock- and SARS-CoV-2-infected blastocysts. Nuclei were stained with Hoechst (white), while NANOG (red) and GATA4 (green) immunostaining identified epiblast and primitive endoderm cells, respectively. Merged images and corresponding single-marker channels are shown. Scale bars, 50 μm in merged image, 20 μm for single channels. **C** Quantification of blastocyst surface area before infection (0 hpi) and during culture following mock treatment (*n* = 47) or SARS-CoV-2 infection (*n* = 48) at 18 and 24 hpi. Multiple unpaired *t* test; adjusted *p*-values are shown. **D** Quantification of total cell number (Hoechst-positive nuclei), NANOG-positive epiblast cells, and GATA4-positive primitive endoderm cells in mock-(*n* = 17) and SARS-CoV-2-infected (*n* = 16) embryos. Mann-Whitney *U* test.

Representative immunofluorescence images of mock- and SARS-CoV-2-infected blastocysts are shown in **Figure 2B**. Nuclei were visualized with Hoechst staining, while NANOG and GATA4 immunostaining identified epiblast and primitive endoderm cells, respectively. Merged images, together with the individual NANOG and GATA4 channels, illustrate the distribution of these lineage-specific markers within the blastocyst.

To determine whether SARS-CoV-2 infection affected blastocyst growth, embryo surface area was monitored throughout the culture period. Before viral exposure (0 hpi), embryo size was comparable between mock and SARS-CoV-2 groups (*n* = 47 mock, *n* = 48 infected). From 18 hpi onward, infected embryos showed significantly reduced surface area relative to mock (18 hpi: *p* = 0.0448; 24 hpi: *p* = 0.0003), indicating that SARS-CoV-2 infection impairs normal blastocyst expansion (**Figure 2C**).

Next, we quantified total cell number and lineage-specific cell populations by immunofluorescence (**Figure 2D**). SARS-CoV-2 infection significantly reduced the total number of Hoechst-positive nuclei compared with mock-treated embryos (*n* = 17 mock, *n* = 16 infected; *p* = 0.0139). This reduction was accompanied by a significant decrease in NANOG-positive epiblast cells (*p* = 0.0054), whereas the number of GATA4-positive primitive endoderm cells remained unchanged (*p* = 0.5255). These findings indicate that SARS-CoV-2 infection compromises blastocyst development by limiting TE expansion and selectively affecting the epiblast lineage while largely sparing primitive endoderm cells.

### SARS-CoV-2 infection induces innate antiviral and inflammatory transcriptional responses in human blastocysts

To characterize the molecular response of human blastocysts to SARS-CoV-2 infection, digital transcriptomic profiling was performed using a NanoString nCounter platform. A total of 57 embryos (29 mock and 28 SARS-CoV-2-infected) originating from five independent experimental cohorts collected as minipools (*n* = 5) of similarly treated embryos were analyzed using a targeted panel comprising more than 750 genes associated with immune responses and viral infection^32,34,35^.

Hierarchical clustering based on the SARS-CoV-2 panel genes indicative of the presence of viral genomic (orf1ab) and antigenomic (orf1ab_rev) as well as subgenomic RNAs (sgRNAs: nucleocapsid, orf7a, envelope, orf3a, and spike) revealed clear expression of viral transcripts in infected embryos from experimental cohorts 1, 2, and 5, whereas embryos from cohorts 3 and 4 showed little or no detectable viral gene expression (**Figure 3A**). While detection of both antigenomic and sgRNA provides unequivocal evidence for successful infection and active viral replication, this pattern indicates variability in infection efficiency between individual cohorts despite identical experimental conditions during inoculation.

**Figure 3.**
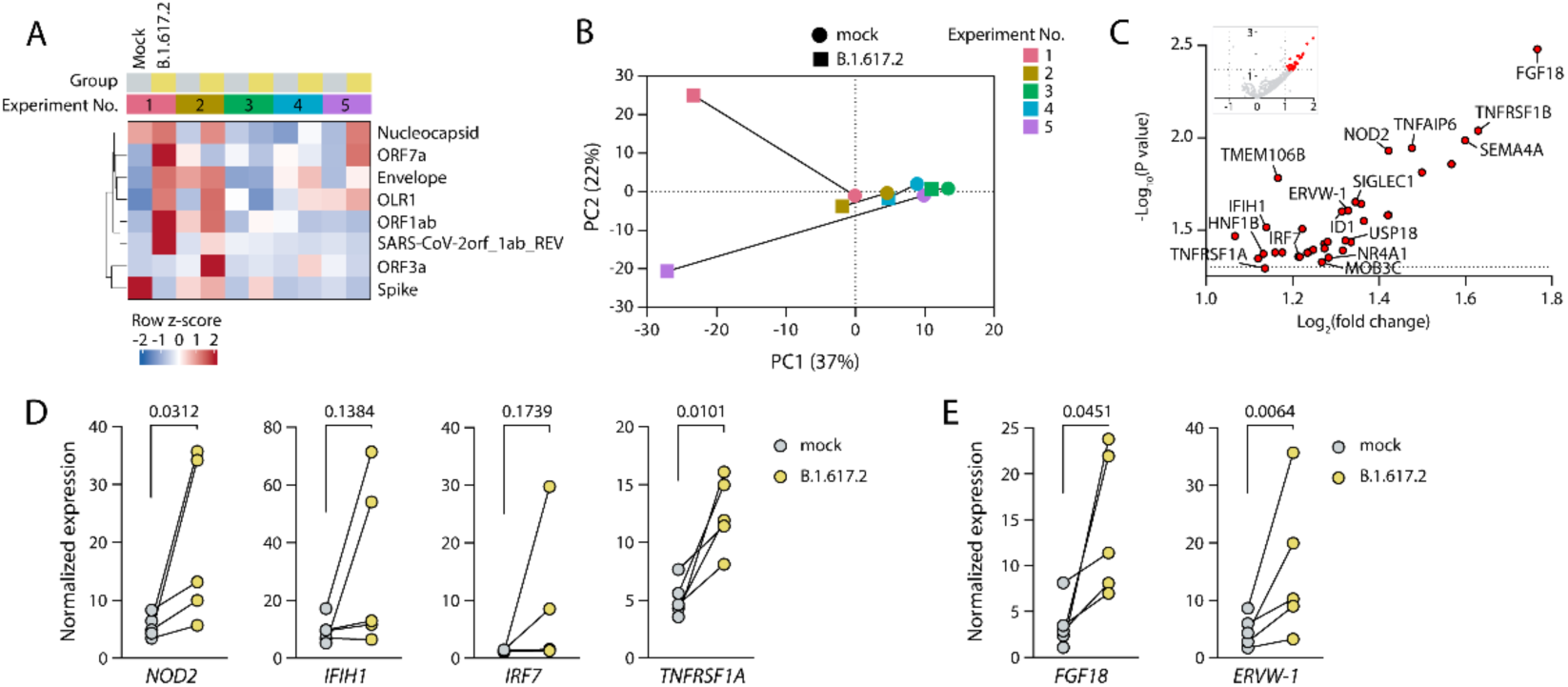
SARS-CoV-2 infection induces innate antiviral and inflammatory transcriptional responses in human blastocysts. **A** Hierarchical clustering heatmap of SARS-CoV-2 viral transcripts detected by NanoString nCounter analysis in mock- and SARS-CoV-2-infected blastocyst pools from five independent experimental cohorts. Each column represents one pool of five to six embryos from the same group and experiment (mock, n = 5 pools, 29 embryos; SARS-CoV-2, n = 5 pools, 28 embryos). Samples are clustered using Euclidean distance and complete linkage. Experimental cohorts are indicated by color annotation. **B** Principal component analysis (PCA) of NanoString transcriptomes based on the complete gene panel (>750 genes). Principal components 1 and 2 explain 37.0% and 22.0% of the total variance, respectively. **C** Volcano plot showing differentially expressed genes between mock- and SARS-CoV-2-infected embryos. Differential expression was analyzed using limma, with experiment and treatment included in the linear model to account for the paired design, followed by Benjamini-Hochberg false discovery rate (FDR) correction. Significantly differentially expressed genes (|log2 fold change| ≥ 1, FDR < 0.05) are highlighted. **D, E** Normalized expression of selected genes involved in antiviral sensing (*NOD2, IFIH1, IRF7*), inflammatory signaling (*TNFRSF1A*), and developmental processes (*FGF18* and *ERVW-1*) in mock (*n* = 5) and SARS-CoV-2-infected (*n* = 5) groups. Each point represents one pool of five to six embryos, and lines connect matched pools from the same experiment. Ratio-paired *t* test.

To investigate global transcriptional changes due to SARS-CoV-2 exposure, principal component analysis (PCA) was performed using all genes included in the NanoString panel. Most mock- and SARS-CoV-2-infected embryos showed substantial overlap, reflecting the overall similarity of transcriptomes. However, infected embryos from experimental cohorts 1 and 5 separated from the remaining samples along the principal components, consistent with their higher abundance of viral transcripts observed in the hierarchical clustering analysis (**Figure 3B**). Together, these findings indicate that the embryos with abundant viral transcript expression displayed the most distinct transcriptional profiles, whereas infected embryos with little or no detectable viral RNA clustered more closely with mock-treated controls.

Differential expression analysis identified a distinct host transcriptional response to SARS-CoV-2 infection (**Figure 3C**). Genes involved in viral RNA sensing and innate antiviral signaling were upregulated in infected embryos, including *IFIH1* (encoding MDA5, a cytosolic sensor of viral double-stranded RNA), *IRF7* (a key transcriptional regulator of type I interferon responses), and *USP18* (an interferon-stimulated gene involved in regulation of interferon signaling)^36–38^. *NOD2*, an intracellular pattern-recognition receptor involved in NF-κB activation^39^, was similarly upregulated. *TMEM106B*, an ACE2-independent SARS-CoV-2 entry factor^40^, was also increased following infection. Infection further induced genes associated with inflammatory signaling, including *TNFRSF1A*, *TNFRSF1B*, and *TNFAIP6*. In addition, several genes linked to developmental processes were differentially expressed, including *FGF18* and *ERVW*-1, which encodes syncytin-1, a mediator of trophoblast fusion and differentiation^41^.

To examine the response of individual genes, the normalized expression of selected genes was compared between matched mock and SARS-CoV-2-infected pools (**Figure 3D, E**). Infection was associated with higher expression of *NOD2* (*p* = 0.0312), *TNFRSF1A* (*p* = 0.0101), *FGF18* (*p* = 0.0451), and *ERVW-1* (*p* = 0.0064). The antiviral genes *IFIH1* and *IRF7* showed a trend toward increased expression that did not reach significance in this paired comparison (*p* = 0.1384 and *p* = 0.1739, respectively).

Together, these findings suggest that SARS-CoV-2 infection is associated with an innate antiviral and inflammatory transcriptional response in human blastocysts, accompanied by trends toward altered expression of genes involved in early embryonic development. The response appeared to be stronger in pools with higher viral transcript abundance than in pools with few viral transcripts, although this was not formally tested.

### Infected embryos show trends toward altered developmental transcriptional programs

To investigate whether SARS-CoV-2 infection influences transcriptional programs associated with early human embryonic development, embryo pool transcriptomes were scored against eight previously defined developmental gene modules representing distinct stages and lineages of preimplantation development, including embryonic genome activation, pluripotency, epigenetic regulation, primitive endoderm, and TE identities (**Figure 4A**)^12^.

**Figure 4.**
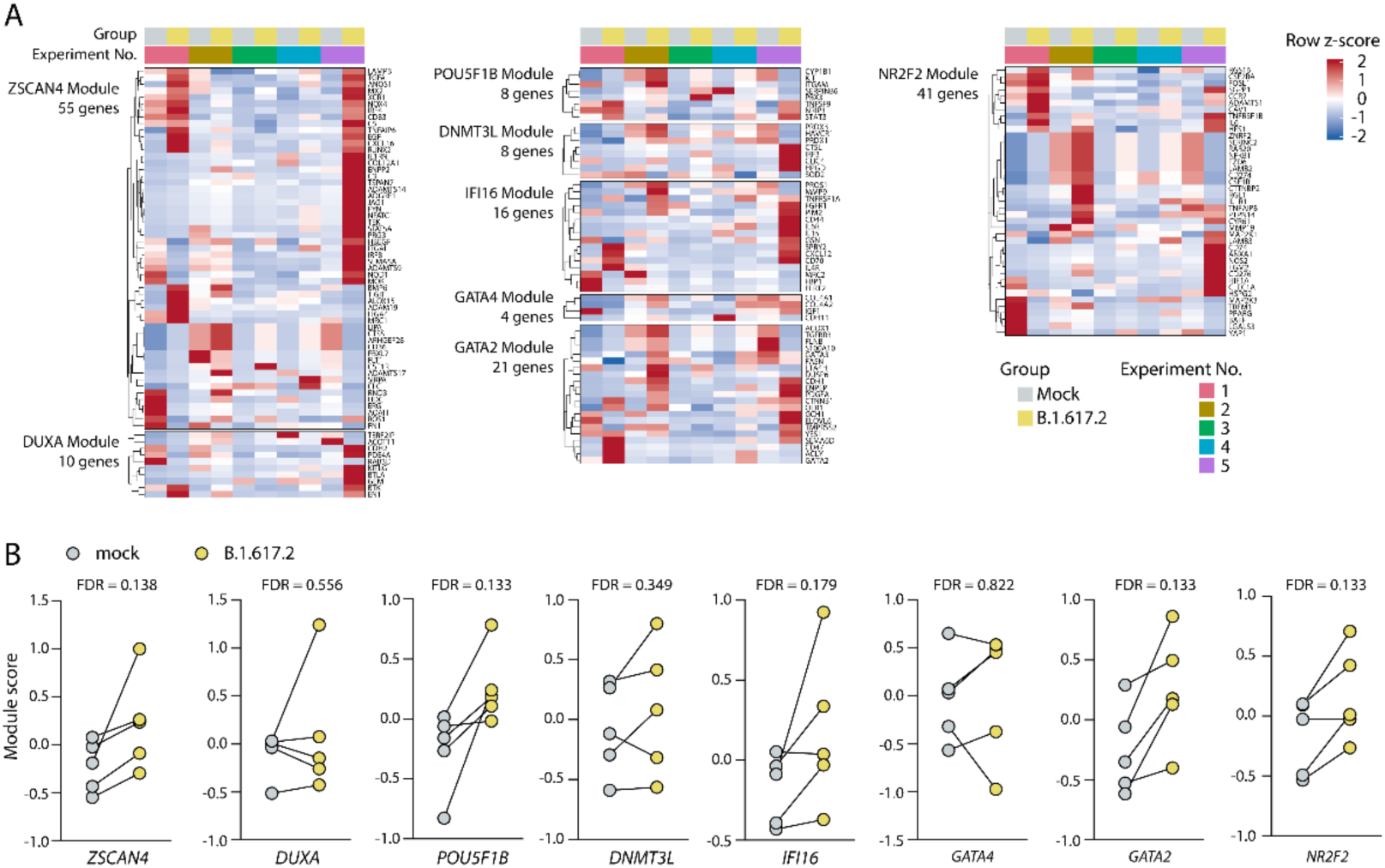
Module analysis suggests coordinated changes in developmental gene programs following SARS-CoV-2 infection. **A** Hierarchical clustering heatmap of developmental gene modules previously defined for human preimplantation embryos ^12^, calculated for each pool of five to six embryos from the nCounter dataset. Modules represent transcriptional programs associated with embryonic genome activation (*ZSCAN4, DUXA*), pluripotency (*POU5F1B*), epigenetic regulation (*DNMT3L, IFI16*), primitive endoderm (*GATA4*), and TE identity (*GATA2* and *NR2F2*). Experimental cohorts are indicated by color annotation. **B** Comparison of module scores between matched mock and SARS-CoV-2-infected pools (n = 5 matched pairs per module). Module scores were calculated as the mean z-score of all informative genes in each module. Two-sided paired *t* tests were performed for each module, followed by Benjamini-Hochberg correction across modules. FDR-adjusted *p*-values are indicated.

Hierarchical clustering of developmental module genes revealed a pattern similar to that observed for the SARS-CoV-2 viral gene panel (**Figure 3A**). Infected pools from experimental cohorts 1, 2, and 5, which exhibited the highest viral transcript abundance, also showed increased expression across multiple developmental modules compared with their matched mock controls, whereas infected embryos from cohorts 3 and 4 clustered more closely with mock-treated embryos (**Figure 4A**).

Module scores were subsequently compared between matched mock- and SARS-CoV-2-infected pools from each experimental cohort (**Figure 4B**). Although none of the eight developmental modules reached statistical significance following correction for multiple testing (all FDR > 0.05), most modules, including those associated with embryonic genome activation *(ZSCAN4* and *DUXA*), pluripotency (*POU5F1B*), epigenetic regulation (*DNMT3L* and *IFI16*), and TE identity (*GATA2* and *NR2F2*), showed higher scores in infected embryos across the majority of experimental cohorts. In contrast, the primitive endoderm module (*GATA4*) showed no consistent difference (FDR = 0.822).

Across several developmental programs, module scores were consistently higher in infected pools, most clearly in the cohorts with the highest viral transcript abundance. None of these differences reached statistical significance after correction for multiple testing, and because transcriptomes were measured in pools of embryos from five matched experiments, the results reflect average responses and should be considered exploratory. Nevertheless, they raise the possibility that SARS-CoV-2 infection is accompanied by shifts in embryonic transcriptional state, and they provide effect-size estimates for designing larger studies. Single-cell RNA sequencing of individual embryos, together with a panel that covers developmental genes, would show which cells and lineages are affected and whether the response is uniform across embryos, and functional analysis of embryo development after infection would test whether these shifts have consequences.

## DISCUSSION AND CONCLUSION

Little is known about the susceptibility, vulnerability and response to viruses of the human embryo. Likewise, SARS-CoV-2 infection of the human embryo during early development represents a major yet understudied concern. While previous experiments using VSV-based viral pseudotypes and engineered reporter viruses suggested that the ZP may protect from viral entry to some extent^17^, our findings indicate that an intact ZP effectively prevents authentic SARS-CoV-2 infection of human blastocysts. Likewise, disruption of this barrier renders embryos susceptible to SARS-CoV-2 infection, even though the underlying TE cells express essential SARS-CoV-2 entry factors such as ACE2, TMPRSS2, and BSG^20,21^. These findings indicate that physical access to embryonic cells, rather than receptor expression alone, determines susceptibility to infection.

Not all established methods of ZP disruption conferred equivalent susceptibility to infection. While LAH consistently resulted in efficient viral infection, TS treatment frequently left residual zona material surrounding the embryo, substantially reducing infection efficiency. This observation is consistent with the bilayered structure of the human ZP, in which the outer layer is more readily dissolved while the compact inner layer is considerably more resistant to acidic digestion^42,43^. Beyond explaining the lower infection rate observed after Tyrode treatment, this offers a practical methodological consideration for future studies of pathogen interactions with human embryos and for investigating differences in the physiological function of individual ZP layers.

Notably, all but one embryo with an intact ZP remained protected. The exception showed viral replication despite apparently preserved zona integrity, but had previously undergone ICSI, suggesting that even submicroscopic discontinuities as created by microneedle puncture during microinjection may suffice to permit viral access, raising the possibility that any experimental procedure compromising ZP integrity may influence susceptibility.

Following ZP disruption, SARS-CoV-2 infection impaired blastocyst development, as evidenced by reduced embryo expansion, decreased total cell number, and a selective reduction in NANOG-positive epiblast cells, while GATA4-positive primitive endoderm cell numbers remained unchanged. The overall reduction in embryo growth is consistent with a generalized developmental impairment, whereas the disproportionate loss of epiblast cells suggests that this lineage may be particularly vulnerable to the downstream consequences of infection. Because viral replication was predominantly observed within the TE cell lineage, these developmental alterations are unlikely to result solely from direct infection of the embryonic lineages themselves but rather from a broader perturbation of the overall developmental program.

A plausible explanation is that SARS-CoV-2 infection disrupts the paracrine communication between the TE and the ICM. Beyond its structural role in blastocyst formation, the TE functions as an important signaling niche for the ICM by secreting factors such as IL-6 that support normal blastocyst development and lineage progression^44^. In addition, TE-derived cells possess a functional type I interferon system capable of protecting embryonic stem cells through paracrine signaling, whereas embryonic stem cells themselves exhibit limited capacity to produce antiviral interferons^45^. Disruption of this supportive relationship by viral infection therefore represents a possible mechanistic link by which TE infection could selectively impair epiblast development without requiring direct viral replication within the epiblast. For example, infection of pre- and peri-implantation embryos with teratogenic Zika virus was largely restricted to the TE yet was sufficient to impair subsequent embryonic and fetal development^46^, supporting the concept that dysfunction of this extraembryonic lineage can influence embryonic development even in the absence of direct infection of the epiblast. This evidence is indirect, as it comes from other models and was not designed to test TE-epiblast signaling, and this mechanism was not tested in the present study; it should therefore be seen as a hypothesis.

Alternative explanations should also be considered. The epiblast may be intrinsically more sensitive than the primitive endoderm to inflammatory stress induced by viral infection. Similarly, the observed phenotype may reflect a generalized developmental delay that disproportionately affects the rapidly expanding epiblast population. The present study cannot distinguish between these possibilities. Future research combining lineage-specific viral detection with single-cell transcriptomics and TE-ICM co-culture systems will be important to determine whether disrupted paracrine signaling represents a direct mechanistic link between TE infection and impaired epiblast development.

Transcriptomic profiling further revealed that SARS-CoV-2 infection induces an intrinsic antiviral response in pools of preimplantation embryos. Increased expression of *IFIH1*, *IRF7*, *USP18*, *NOD2*, and TNF-associated signaling pathways indicates that embryonic cells can detect viral infection and activate innate immune programs before implantation, consistent with previous evidence that human preimplantation embryos express functional pattern-recognition receptors and respond to pathogen-associated stimuli^47^. The increased expression of *TMEM106B*, recently proposed as an ACE2-independent host factor for SARS-CoV-2 entry^48,49^, raises the possibility that alternative viral entry mechanisms may contribute to embryonic susceptibility, warranting further investigation.

The extent of transcriptional responses varied largely between individual embryo cohorts, with two cohorts showing stronger induction of antiviral as well as interference with developmental gene modules than the others. Obviously, the observed variability may reflect variation in infection efficiency. However, more likely, differences in embryo quality, such as donor-specific characteristics and an experimentally more difficult-to-control timing of developmental stages in the already heterogeneous pools, can be expected to have a greater impact. Indeed, future analysis of established developmental gene clusters in mock samples can be expected to assign individual cohorts to at least three different stages on the earlier established pseudotime^12^. Only five experiments were analyzed, and embryos had to be pooled because a single embryo does not yield enough RNA for detection, so individual embryos could not be studied; our conclusions are therefore preliminary. The variability we observed between experiments may partly reflect the natural biological variability of human embryos, which should be kept in mind when interpreting responses to viral infection.

While our experimental model demonstrates that human blastocysts can support SARS-CoV-2 infection as soon as the ZP barrier is compromised, the relevance of this scenario to natural conception requires careful consideration. Additional physiological barriers can be expected to limit infection of the embryo during natural conception. The intact ZP physically isolates the preimplantation embryo during transit through the reproductive tract, and productive SARS-CoV-2 infection of the female reproductive tract itself appears uncommon. Indeed, viral mRNA was undetectable in endometrial biopsies from women with active COVID-19 despite ACE2 expression in this tissue^50^. Although SARS-CoV-2 RNA has been sporadically detected in semen and reproductive tissues, studies in acute COVID-19 have generally failed to demonstrate consistent seminal shedding^51^. Detection of viral RNA in semen from some individuals with Long COVID has raised the possibility of persistent reproductive tract reservoirs^52^. However, whether this represents infectious virus with implications for fertility and transmission remains unresolved.

Embryonic infection *in vivo* would additionally require a tightly synchronized sequence of events, including maternal infection during the narrow preimplantation window and sufficient viral dissemination to the uterine environment before implantation. Direct embryonic infection during natural conception is therefore likely rare; adverse reproductive outcomes associated with maternal COVID-19 more plausibly arise through indirect mechanisms, including systemic inflammation, cytokine dysregulation, altered endometrial receptivity, or placental dysfunction after implantation^53–55^. Maternal immune activation may represent an additional mechanism by which infection can influence early development. Although a controlled inflammatory response is an integral part of implantation and early pregnancy, excessive or dysregulated production of inflammatory mediators can disrupt the cellular and molecular processes required for normal embryonic development^56,57^. Thus, even in the absence of direct embryonic infection, maternal infection-associated inflammation may create an adverse developmental environment during critical windows of early pregnancy.

The implications may differ in MAR settings, where procedures such as LAH or ICSI intentionally breach the ZP. Laboratory transmission of SARS-CoV-2 is considered extremely unlikely under modern biosafety protocols^58,59^, but our findings show that embryos become susceptible once this physical barrier is compromised. The viral inoculum used here was chosen to establish a reliable experimental infection and does not reflect typical clinical exposure levels; the consequences of lower-dose exposure, and whether limited viral replication could influence implantation, lineage specification, or later fetal development, remain open questions. Importantly, these findings do not suggest that routine IVF procedures pose a substantial risk of SARS-CoV-2 transmission, or viral infections in general. Nevertheless, they highlight the importance of understanding how specific micromanipulation procedures may alter the physical barriers that normally protect the embryo.

Several experiments could validate our main conclusions. First, repeating the single-cell RNA sequencing approach used to define the developmental modules^12^ on infected and mock embryos would show which cells and lineages respond, and whether the response is uniform across embryos. Second, following individual embryos, with measurement of their development, lineage composition, and gene expression, would link infection directly to fate, which pooling did not allow. Third, larger cohorts, other viral variants, and quantitative measurement of infection in each embryo would test whether the response depends on the amount of virus.

### Limitations of the study

Several limitations should be considered when interpreting our findings. First, our study evaluated a single SARS-CoV-2 variant, Delta (B.1.617.2), and viral entry or replication characteristics may differ between variants. However, the latter variant may serve as a representative prototype, given that the Delta variant, which became the dominant global strain by mid-to-late 2021 and drove a surge in infections, exhibited a particularly strong impact on reproductive health^6^. Moreover, the direct and timed exposure of ZP-disrupted embryos to a defined viral inoculum does not recapitulate the complex conditions of natural conception. The *in vitro* system further lacks maternal immune, endocrine, and uterine interactions that shape early pregnancy. Either contribution will need to be studied in future experiments, for which we herewith provide a base dataset. In addition, infection was confirmed by dsRNA staining, which indicates viral replication but does not quantify the level of infection per embryo. Second, digital transcriptomics (nCounter) is a targeted method. The panel used here focuses on innate immunity and was not designed for early embryonic or pregnancy development, so genes relevant to these processes may have been missed, and novel transcripts or pathways cannot be identified. Its sensitivity, of digital transcriptomics, though possibly equally, if not more, sensitive than other methodologies such as RNA sequencing^60^ is also limited. In combination with the low cell number of human preimplantation blastocysts at 6 dpf (∼200)^61^ and resulting low RNA yields, this required pooling of individual embryos. While use of minipools may have helped average experimental errors between biological replicates, a striking inter-cohort variability remains. Likewise, the limited number of pools available for transcriptomic analysis resulted in reduced statistical power for exploratory pathway and developmental module analyses; therefore, these findings require validation in larger cohorts. Nevertheless, disruption of certain transcriptional patterns linked to an established pseudotime of early development suggests a risk of loss of lineage commitment and stage-specific vulnerabilities^12^. However, pinpointing such specific vulnerabilities should be feasible from transcriptional data sets as presented herein. Third, the embryos were surplus embryos donated to research, and no clinical background information on the donors, such as maternal conditions or the indication for PGT, was available, and the sex of the embryos was unknown. These factors could therefore not be taken into account. Random allocation of embryos to matched mock and infected groups within each experiment was used so that such variability would be distributed across both groups. Finally, although our data support a model in which TE infection indirectly affects epiblast development, the underlying mechanisms remain to be established. Targeted disruption of developmental programs, such as by viral infection in the current study, may help elucidate such natural dependencies. The *in vitro* system also cannot capture maternal factors or assess longer-term consequences after implantation, which will require further investigation.

## Conclusion

In conclusion, our findings identify the human ZP as the principal barrier preventing SARS-CoV-2 infection of preimplantation embryos. Once this barrier is disrupted, authentic SARS-CoV-2 readily infects blastocysts, impairs embryonic growth, selectively affects epiblast development, and induces a cell-intrinsic antiviral response. While direct embryonic infection during natural conception is likely uncommon, these findings provide mechanistic insight into early embryonic susceptibility to viral infection and underscore the importance of maintaining appropriate biosafety considerations during MAR procedures.

## Data Availability

Datasets generated and/or analyzed during the current study are provided in the paper or are appended as supplementary data. Source data are available upon reasonable request to the authors.

## Acknowledgments

The authors are grateful to Elisabeth Heylen for her advice in the execution of experiments under controlled biosafety level 3 conditions; to UZ Brussels, in particular, the patients who kindly donated embryos for this research; and Yves Guns, Sabrina Vitrier, and all the other Brussels IVF personnel, for facilitating our research. And the funding agencies FWO and the European Union for making this research possible.

## CRediT Authorship Contribution Statement

Conceptualization, W.E., Y.A.A., K.D., H.V.V.

Data curation, Y.K., J.V.W., Y.A.A.

Formal analysis, Y.K., J.V.W., Y.A.A.

Funding acquisition, Y.K., W.E., Y.A.A., K.D., H.V.V.

Investigation, Y.K., M.T., W.E., Y.A.A.

Methodology, Y.K., M.T., W.E., Y.A.A., K.D., H.V.V.

Project administration, Y.K., Y.A.A., K.D., H.V.V.

Resources, N.D.M., J.V.W., K.D., H.V.V.

Supervision, Y.A.A., K.D., H.V.V.

Validation, Y.A.A., K.D., H.V.V.

Visualization, Y.K., Y.A.A.

Writing – original draft, Y.K., Y.A.A., K.D., H.V.V.

Writing – review & editing, all authors.

All authors have read and agreed to the published version of the manuscript.

## Declaration of generative AI and AI-assisted technologies in the writing process

Statement: During the preparation of this work, the authors used ChatGPT and Claude to improve the readability and language of the manuscript. After using this tool/service, the authors reviewed and edited the content as needed and take full responsibility for the content of the published article.

