## Supplementary Figure 1 for "SARS-CoV-2 susceptibility and vulnerability of human blastocysts during *in vitro* preimplantation development"

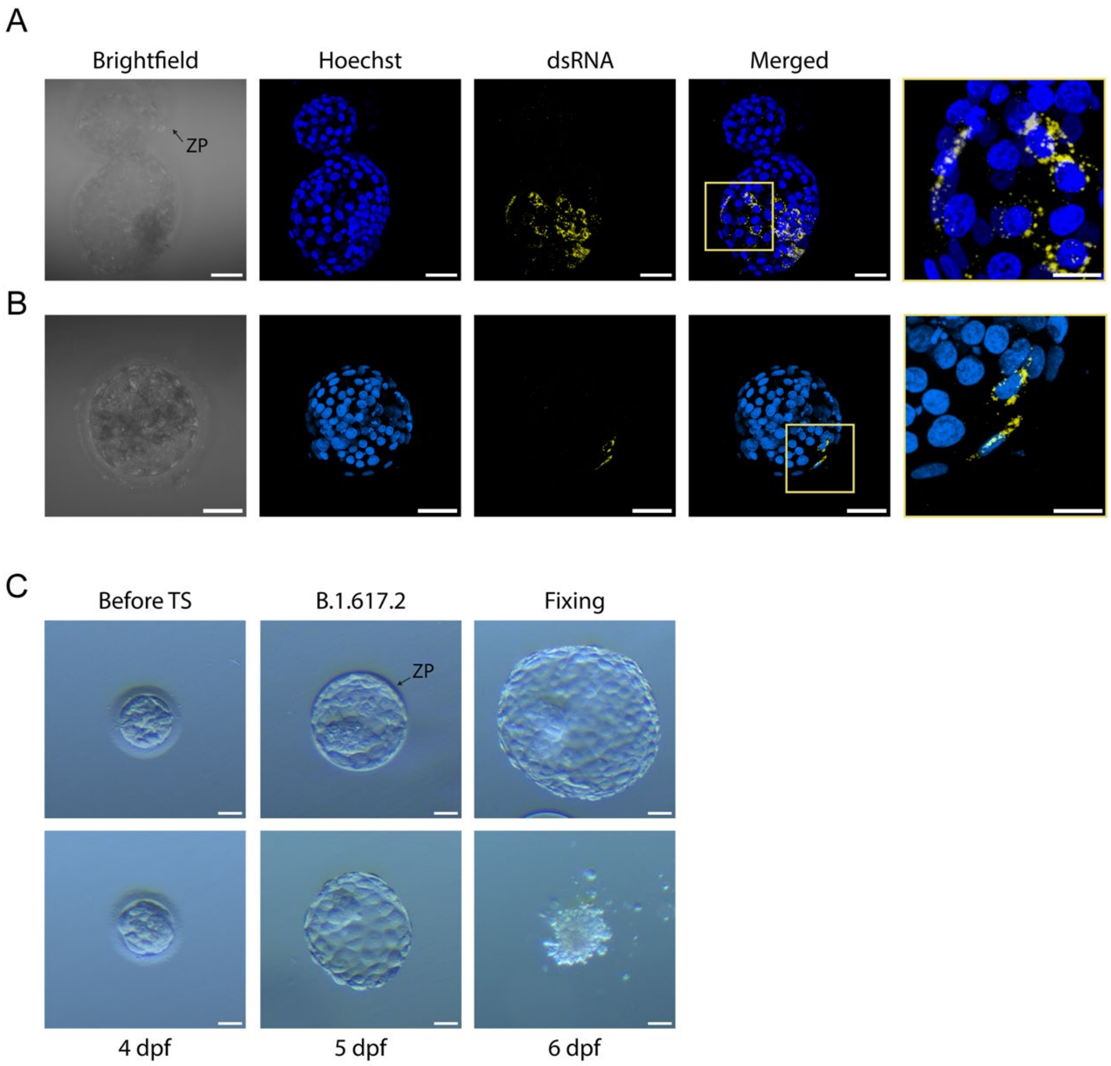


**Supplementary Figure 1. Additional observations following SARS-CoV-2 infection of human blastocysts. A, B** Representative immunofluorescence images of SARS-CoV-2-exposed blastocysts with an apparently hatching region (A) and an intact zona pellucida (ZP) (B), both positive for double-stranded RNA (dsRNA, yellow), indicative of viral replication. Nuclei were stained with Hoechst (blue). Scale bars, 50 µm; 20 µm in the magnified images. **C** Representative brightfield images of Tyrode's solution (TS)-treated embryos following exposure to $3x10^4$ $TCID_{50}$/ml SARS-CoV-2. The upper panel shows an embryo with residual ZP remaining after TS treatment that retained a morphologically intact appearance. The lower panel shows an embryo in which the ZP was completely removed; following viral exposure, the embryo exhibited extensive degeneration and lysis. Scale bars, 50 µm.
